# HIV-HBV Co-infections in Central India: Prevalence, Clearance Rates, and Long-term Outcomes from a Decade of Experience at a Tertiary Care Center

**DOI:** 10.1101/2023.10.05.23296399

**Authors:** Uppal Rashmi Kunkal, Nitu Mishra, Jyoti Tiwari, Talha Saad, Ashish Kumar Vyas, Amardeep Rai, Sumit K Rawat

**Affiliations:** Department of Microbiology, Bundelkhand Medical College Sagar, Madhya Pradesh, India; Department of Medicine, Bundelkhand Medical College Sagar, Madhya Pradesh, India; Department of Pulmonary Medicine, Bundelkhand Medical College Sagar, Madhya Pradesh, India; Dept. of Immunology, John C Martin centre for Liver Research, Kolkata, India

**Keywords:** HIV, Antiretroviral Therapies, Vaccines, HIV transmission, MSM

## Abstract

Two of the most infamous viruses, the Human immunodeficiency virus (HIV) and hepatitis B virus (HBV) have similar transmission routes. Consequently, some individuals might get co-infected with both viruses resulting in increased deaths and prolonged disease. Even among healthy (Non-immunocompromised) persons, chances of getting persistent HBV infection are 5-10%, however among persons harboring HIV, HBV the persistence rates might escalate up to 15 %. Such increased suffering might occur despite the longevity provided by starting highly active anti-retroviral therapy (HAART), which is the current standard treatment.

Our pioneering study, in our region aims to assess the prevalence of HIV-HBV co-infection, clearance rates of HBV, demographic characteristics of affected individuals, and long-term outcomes. By shedding light on these aspects, we hope to gain valuable insights into the impact of such co-infections and pave the way for better management and care of individuals facing this dual challenge.

We studied 1808 persons enrolled for HIV treatment from amongst 170,019 persons screened. Higher co-infection of HIV-HBV was observed in males as compared to females, with the age group of 31-40 years being most affected. The most common route of infection, was heterosexual contact accounting for 86% of cases, accounting for majority co-infections. Over the years, we noticed a decline in the number of HIV-HBV co-infected cases, with the nadir occurring in 2015. Seroprevalence of co-infections was 2.37%, but despite this observed low sero-prevalence HBV clearance was relatively poor among the co-infected, as only 4 (9.3%) able to clear the infection after initiation of HAART. Our study highlights that chances of HBV clearance among the co-infected are worse than that observed among otherwise heathy populations thus also highlighting the urgent need for universal HBV vaccination in all HIV-affected persons, underscoring the importance of providing special attention to them.

## Introduction

The Human immunodeficiency virus (HIV) is one of the most dreaded infections of mankind with a global burden of 39 million affected people, total 63 thousand attributable deaths and 1.3 million new infections in 2022 (1). The global response to HIV has made significant progress in recent years, it has been well documented that after the treatment started with highly active antiretroviral therapy (HAART), persons living with HIV are living longer and experiencing lesser health problems. With the advent of this treatment, HIV has been given a status of chronic disease and those who acquire these infections in a young age will live long enough either to experience morbidity due to diseases other than acquired immune-deficiency syndrome (AIDS), like liver diseases, chronic neurological problems or they might acquire new infections such as HBV, hepatitis C virus (HCV) or hepatitis D virus (HDV) in this prolonged life span (2).

HIV and HBV besides sharing similar transmission routes are known to be associated with increased morbidity and mortality in persons having their co-infections (3). Among such infections, chronic HBV infection continues to be a global public health problem with nearly 240 million chronically infected with HBV, with India and other Asian countries having one of the highest prevalence of chronic HBV infection. Among healthy subjects due to spontaneous HBV clearance chances of getting a chronic infection of HBV are 5-10%.

However, in the persons living with HIV the chances of HBV clearance are very low and most of the HBV infections become persistent which is often denoted as chronic HBV (4). According to the current knowledge, the chances of getting co- or super-infections by other microorganisms among them still remain due to the impaired host immunity, such co-infections including HBV in such persons is not only associated with increased suffering but sometimes leads to a worse outcome (5). Studies have found that due to HIV immune suppression, asymptomatic careers may also lead to active hepatitis and high HBV viral loads because of HBV reactivation. In such individuals, HIV interferes with the natural history of HBV infection through enhancing HBV replication which could lead to more severe and chronic liver disease (6). Such chronic liver infections are known to directly impact hospitalizations and mortality among the affected (7).

Approximately, 600,000 people die each year with HBV-related liver diseases, this becomes more important in persons affected with HIV since HBV co-infection might negatively impact outcomes among those already harboring HIV. In comparison to healthy individuals, those co-infected with HIV-HBV are 17 times more likely to die of liver-related causes (2). These considerations made the Centers for Disease Control (CDC) to make a powerful argument for the importance of HBV vaccination for all individuals with HIV infection (2); However, no such recommendations or practices are being followed despite studies reporting that these high-risk populations have a very low HBV vaccine coverage in developing countries like India (8).

A whopping number of 686000 people die in India every year as a consequence of HBV infection(9). In a recent study around 1% of children in India below 14 years are found to be chronic HBV carriers despite the availability of an effective vaccine and the inclusion of HBV vaccination in the immunization schedule (10). Having poor vaccination coverage India alone accounts for around 40 million HBV carriers, of whom 15% to 25% develop cirrhosis and various complications leading to increased healthcare costs and adverse outcomes (11).

It is imperative to know the factors which play a role not only in the overall prevalence of HIV-HBV co-infection but also in the disease pattern and the fate among the affected. Prevalence and fate of HIV-HBV co-infection depends upon multiple factors such as immunization status, HIV viral load, route of transmission, immune or nutritional status and the time of initiating HAART, etc. In western countries, they have a high HIV-HBV co-transmission leading to high seroprevalence (12), leading up to a reported prevalence of HIV-HBV co-infection as high as 9-12%. It is attributed to practices such as intravenous drug use and male-to-male sexual contact, which facilitate more effective viral transmission (3). Similarly, studies from the eastern part of India have found a high prevalence of HIV-HBV co-infection especially among persons using the intravenous drugs and MSM population (13). Contrarily, regions in the rest of India, where infection is primarily acquired through heterosexual routes, tend to exhibit lower prevalence rates (14).

India’s vastness, socio-economic disparities and highest population burden in the world call for region-specific studies. On exploring such region-wise published data from India, HIV-HBV co-infection rates studied are mostly from urban areas, these studies come from selected regions mainly from Eastern India or Northern India (15), some are from South India, and very few studies from the Western part of India (14,16).

Unfortunately, little attention has been given to the central Bundelkhand region, which is a large area affected by undernutrition, poor health coverage, and poverty. Literature is lacking regarding the prevalence of HIV-HBV co-infection, its route of acquisition, the clearance rates and the outcome of HBV among persons living with HIV. As one the largest tertiary care institute in such underprivileged area, serving a substantial tribal population, we aim to investigate HIV-HBV co-infection rates, shedding light on this unexplored aspect and contributing to better healthcare outcomes.

## Materials & Methods

### Population

The current study is a retrospective database review and follow-up record analysis of individuals diagnosed with HIV and enrolled for Highly Active Antiretroviral Therapy (HAART) at a tertiary care center in Central India, spanning from October 2010 to December 2020. The cases were diagnosed from a pool of 170,019 persons tested for HIV at our institute, which also serves as a referral center for neighboring areas and districts in central India.

### Inclusion criteria

All subjects who tested positive for HIV and fulfilled the requirements for HAART initiation, subsequently enrolling for treatment.

### Exclusion criteria

Patients deemed unsuitable for HAART or those who did not complete a single follow-up visit.

As per the National AIDS Control Organization (NACO), enrolment protocols followed at the ART center as shown in the process flow diagram in figure 1. Blood and serum samples for routine screening were obtained for testing/monitoring from each patient after confirmation/detection of HIV status at baseline and on each follow-up visit (17).

**Figure 1:**
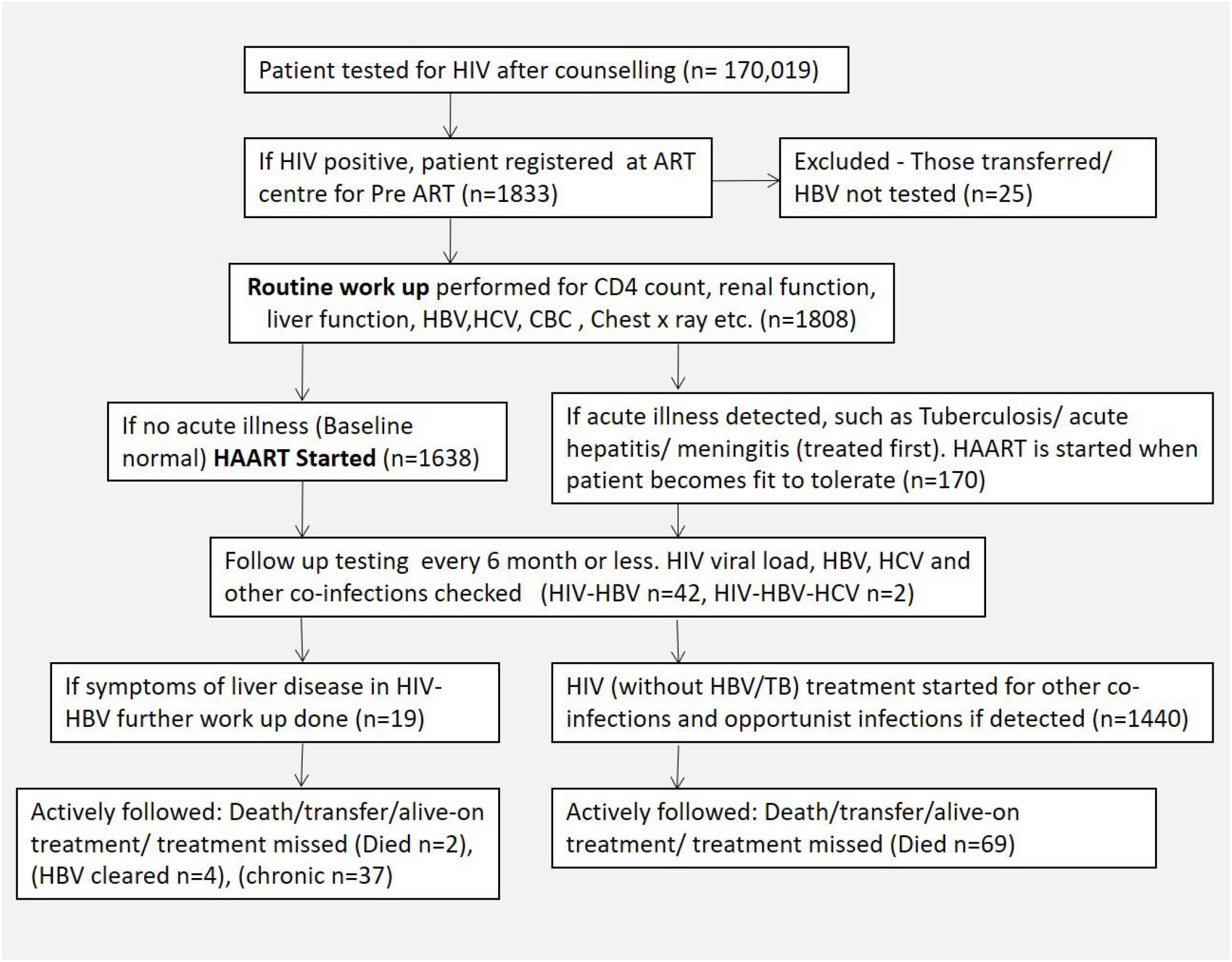
Protocol for enrollment and work-up of the study population at ART center.

Various specimens EDTA whole blood, serum and plasma from these patients were subjected to workup including as mentioned below

### Serology and virology

HIV-1/2 antibody testing was performed by rapid spot-based assay as per NACO protocols (Parikshak, Tri-dot and comb HIV). Hepatitis B surface antigen (HBsAg) and Hepatitis C antibody testing was performed by SD Bioline rapid immunochromatographic assay (17).

Samples tested reactive for HBsAg by immuno-chromatographic test, were reconfirmed by commercial ELISA kits using freshly redrawn samples (Microlisa J Mitra & Co PVT LTD). Virology testing was available from 2014 onwards, where HIV viral load was performed by quantitative real-time qRT-PCR using Roche COBAS® AmpliPrep/COBAS® TaqMan® HIV-1 Test, while HBV viral load, was performed by real-time cassette based PCR TRUENAT HBV (Molbio India).

Sputum AFB testing at OPD visits along-with chest X-ray and Genexpert MtbRif testing of sputum/body fluid for tuberculosis was performed to rule out co-existing tuberculosis (18).

### Biochemical tests

Liver function tests namely alanine aminotransferase (ALT), aspartate aminotransferase (AST), alkaline phosphatase (ALK), serum albumin, serum bilirubin, and kidney function tests (Urea, creatinine, uric acid, serum electrolytes) were performed in the department of Biochemistry by Biosystem BA-400 fully automated system with supplied kits and as per manufacturer instructions.

### CD4 and CD8 Quantification

CD4+ counts were performed for all the patients by BD FACS Count system on EDTA whole blood, which was collected in a vacutainer, maintaining aseptic precautions at the ART center, and then processing was performed as per manufacturer protocols in the department of microbiology.

### Statistical analysis

Data were extracted and entered into MS Excel, and proportions/percentages were calculated for categorical variables. The chi-square test was used to examine possible associations between categorical variables such as seropositive HBV-HIV co-infected patients versus HIV only, considering gender. For variables with normal distribution, a t-test was applied to compare mean values of different baseline parameters like age, weight, CD4 count on presentation, and platelet count. The Mann Whitney U test was used for variables without normal distribution. A p-value of less than 0.05 was considered significant. The statistical analysis was performed using SPSS trial version 16 for Windows.

A total of 125 Patients who presented with HIV-TB co-infection were excluded from the statistical analysis among the HBV-negative comparison group to prevent co existing tuberculosis infection as a confounder. No such exclusion was required among HIV-HBV co-infected patients, as none of these tested positive for tuberculosis at presentation.

Ethics statement: The review and analysis work was performed after obtaining due approval from the institutional human ethics committee.

## Results

We found that 43 individuals were co-infected with HIV and HBV with a sero-prevalence of 2.37% amongst the total 1808 who could be tested for HBsAg after registration for Pre-ART/HAART among total 1833 individuals who had presented to the ART center for receiving treatment. It is pertinent to note that total 25 persons were excluded from the study as they could not be tested for HBV infection as either they died before work-up or they were transferred other institutes.

Our study encompassed a wide age spectrum, ranging from neonates to individuals as old as 85 years, with the average age at presentation being 36.2 ± 10.8 years. Figure 2 illustrates the distribution of co-infected patients across various age groups over the years.

**Figure 2:**
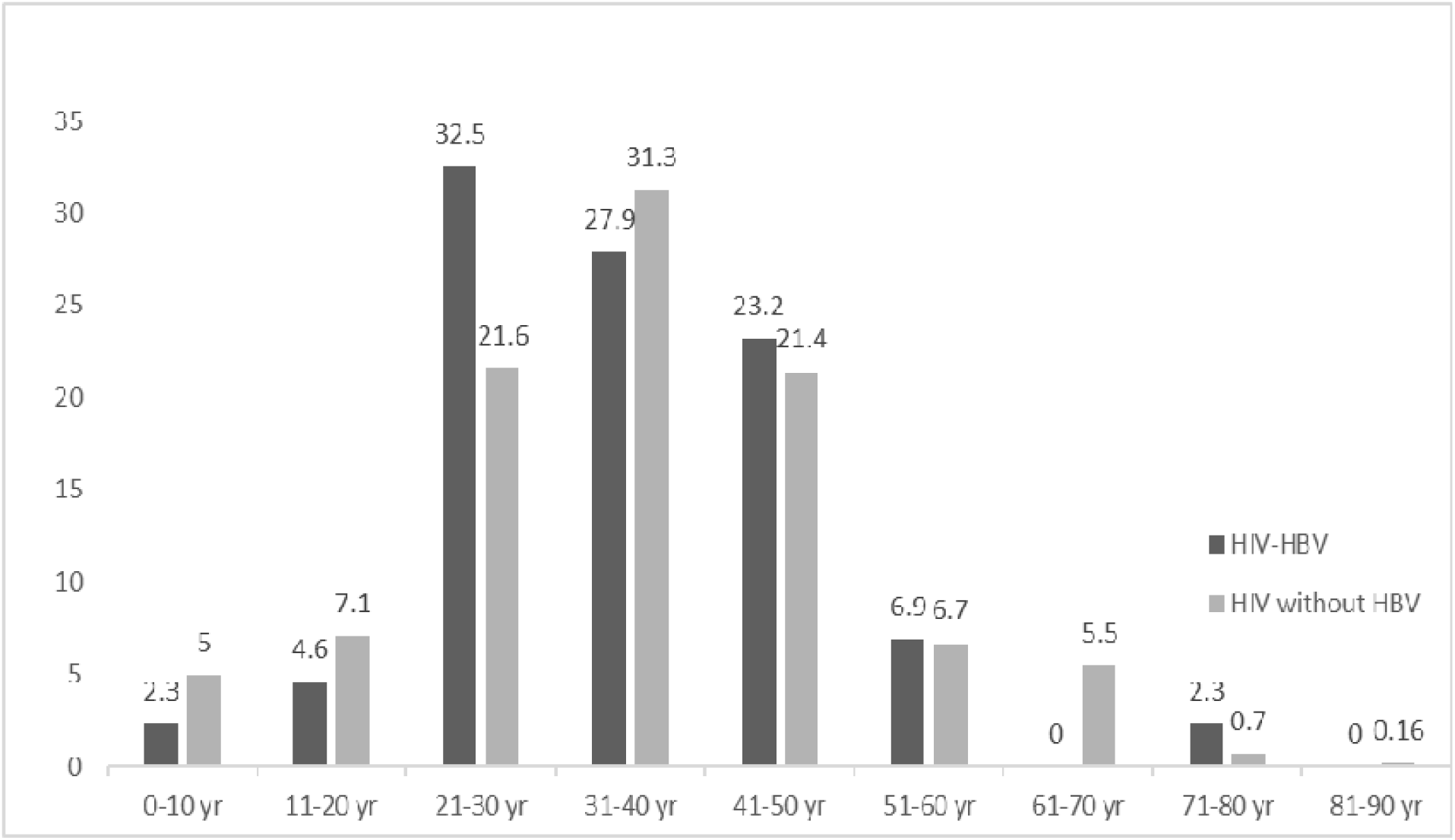
Age-wise distribution of study population at presentation.

The study population had male predominance with 1126 (62.27%) males inclusive of 1063 adult males, 63 male children, 680 (37.61%) females inclusive of 631 adult females, 49 female children, and 2 (0.11%) adult transgender (Table 1). As regards to the mode of acquisition of HIV-HBV coinfection as shown in figure 3, the commonest HIV transmission route observed in our study was heterosexual, which was seen in 37 (86%), and transmission through drug injections was least common. The route of infection could not be ascertained in one of the co-infected persons and was unknown.

**Figure 3:**
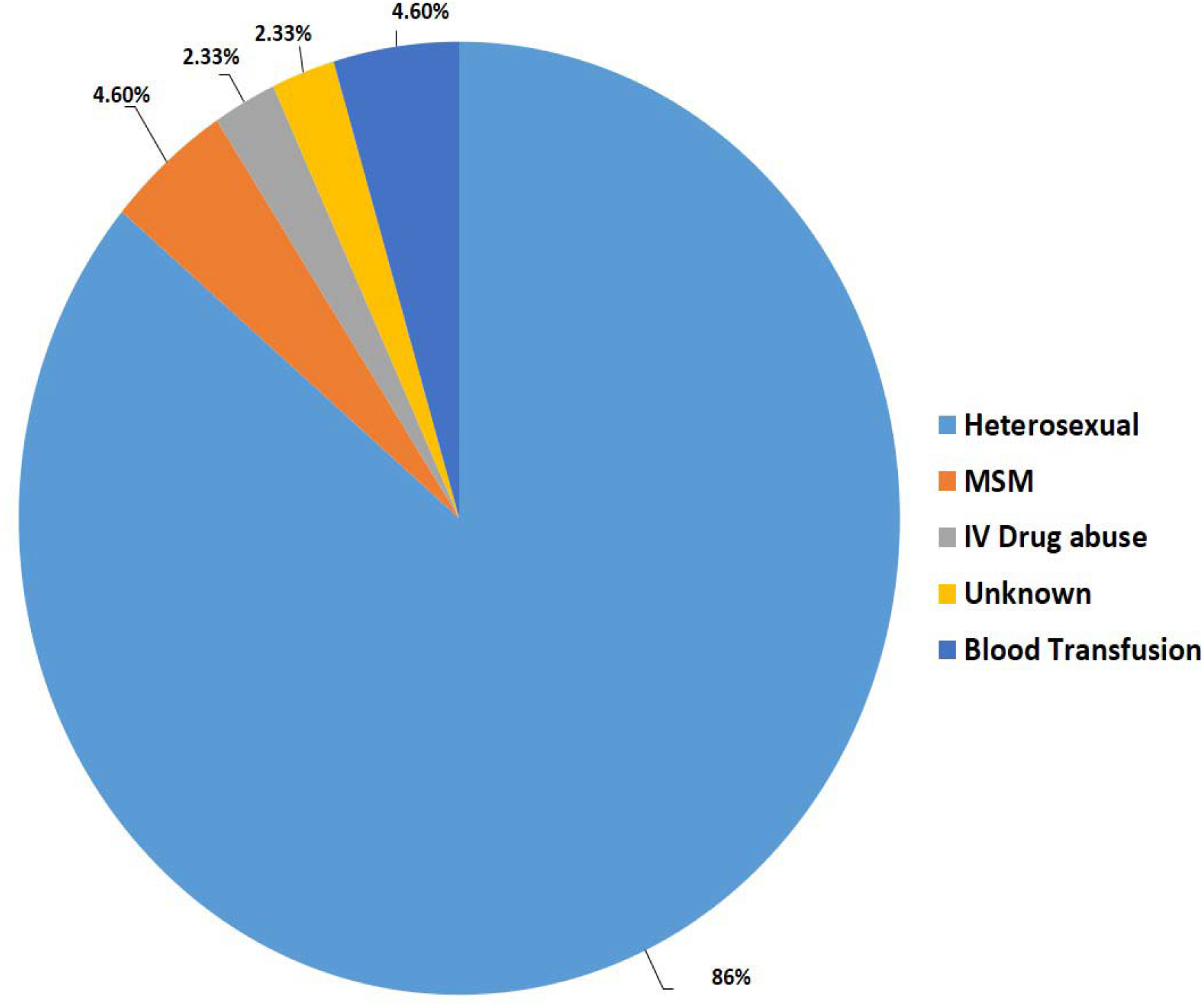
Modes of acquisition of infection among the HIV-HBV co-infected.

**Table 1:** Age and sex distribution of study group versus the subset of HIV-HBV co-infected patients.

|  | Adult males | Adult females | Male children | Female children | Transgender |
| --- | --- | --- | --- | --- | --- |
| Total patients<br>(n=1808) | 1063(58.7%) | 631(34.9%) | 63 (3.5%) | 49 (2.7%) | 2 (0.11%) |
| HBV-HIV co-infected (n=43) | 24(55.8%)<br>*2 patients<br>HBV/HCV/HIV co-infected | 16(37.2%) | 2(0.46%) | 1(0.23%) | 0 (0 %) |

The observed year-wise distribution of HIV-HBV co-infection cases was highest in 2015 (n=8 in males, n=4 in Females) with the lowest cases in 2020 (n=1 male). In our study, the maximum number of cases registered were in the age group of 31-40 years followed by those of 21-30 years.

The mean values of various baseline characters on presentation in both the groups of patients along with their obtained statistical significance values are shown in Table 2. Among HIV only group a total 45 showed signs of liver dysfunction, however, these signs were mostly drug-induced and subsided on substituting the respective anti-retroviral drug.

**Table 2:** HBV surface antigen status, baseline demographic and laboratory data of study population.

|  | HBV surface antigen status |  | *P value |
| --- | --- | --- | --- |
|  | HIV-HBV co-infected<br>(n=43) | HIV Positive Only<br>(n=1640) |  |
| Male sex, n (%) | 26 (60.4) | 964 (58.8) | 0.92 |
| Age at diagnosis (Yrs.) | 36.16 ± 12.39 | 36.64 ± 10.47 | 0.81 |
| AST (U/litre) | 46.07 ± 29.78 | 35.60 ± 20.01 | 0.13 |
| ALT (U/litre) | 41.06 ± 26.01 | 37.85 ± 24.22 | 0.58 |
| Creatinine (mg/dl) | 0.96 ± 2387.37 | 1.02 ± 0.33 | 0.30 |
| TLC (/mm<sup>3</sup>) | 6515.92 ± 2387.37 | 7348.03 ± 3009.09 | 0.13 |
| CD4 count (/mm<sup>3</sup>) | 322.63 ± 229.49 | 219.64 ± 157.38 | 0.02 |
| Average weight (kg) | 49.5 ± 12.13 | 45.48 ± 10.95 | 0.12 |
| Platelet count (/mm<sup>3</sup> X 10<sup>3</sup>) | 1.93 ± 0.73 | 2.34 ± 0.71 | 0.12 |
| Died, n (%) | 2 (0.46) | 69 (0.42) | 0.87 |
| *Mann Whitney U test wherever means are compared and Chi square test for discrete data. |  |  |  |

**Table 3:** Various parameters amongst those with HBV persistence versus those with HBV clearance.

|  | HBV persistence (n=39) | HBV cleared (n=4) | *P value |
| --- | --- | --- | --- |
| HbeAg | 5/15 | -- | -- |
| Hbe Ab | 2/15 | 3/4 | -- |
| Anti-Hbc Ab | 0/15 | 4/4 | -- |
| <sup>\$</sup> Anti-HCV | 2/39 | 0/4 | -- |
| Anti-HDV | 0/39 | 0/4 | -- |
| Age (Yrs.) | 36.43 ± 12.59 | 33.5 ± 11.38 | 0.58 |
| CD4 mean ± SD | 314.65 ± 232.8 | 374.5 ± 230.5 | 0.36 |
| <sup>\$</sup> HBV/HCV/HIV co-infected, HDV/HBV/HIV co-infected.<br>*Mann Whitney U test | | | |

Among the HIV-HBV co-infected the mean CD4 counts were higher than the mean CD4 counts of HBV negative group. The MSM sub-group in the HIV-HBV had far lower CD4 counts and had an advanced disease staging as compared to those with other modes of acquisition. Mean values of liver enzymes were higher than the HBV negative cases, with 5 of them having marked derangements (>2x upper normal limits) in liver function. They had complaints of loss of appetite, vomiting, and other signs of liver dysfunction. Two of them had CD4 counts below 500/cu mm and were found to have fatty infiltration with hepatomegaly on ultrasound, the rest 3 of them, fortunately had CD4 counts >500/cu mm. Among these upon further work-up like liver ultrasound, hepatitis B envelope antigen (HBeAg), anti-hepatitis B core antibodies (anti-HBcAb), etc. HBeAg was positive in all 5 patients, none of them had developed anti-HBcAb.

Follow-up and outcome: Among those who ever presented to ART center, many patients didn’t turn up for follow-up over the years and it was noted that overall 260 patients had died, 64 of these could not receive any form of ART. During the study period, a total of 1638 patients could satisfy the prerequisite to be put on HAART, the remaining 170 could register only for pre-ART. Among the study group 196 (10.8%) died due to various causes, the most common of them being HIV-TB co-infections accounted for 125 (6.9%) deaths, followed by anemia, diarrhea, pneumonia and hepatitis.

Among those having HIV-HBV 2 (0.86%) died, both males, having CD4 counts below 500/cu mm. One of these was an adult who had chronic hepatitis with cirrhosis and died at 1 year of diagnosis after going into hepatic encephalopathy, the second was a child who was underweight for his age and died within 6 months of registration. Besides HBV co-infection, none of these had any other obvious reasons leading to death.

Amongst the other group (HIV only) total of 69 patients died (Tb co-infection excluded) and with one death due to a liver-related cause (alcoholic liver cirrhosis). HIV-HBV co-infection not only had higher percentage mortality (table 2), but the mean years of survival among the co-infected were also less than that of persons living with HIV only as shown in figure 4.

**Figure 4:**
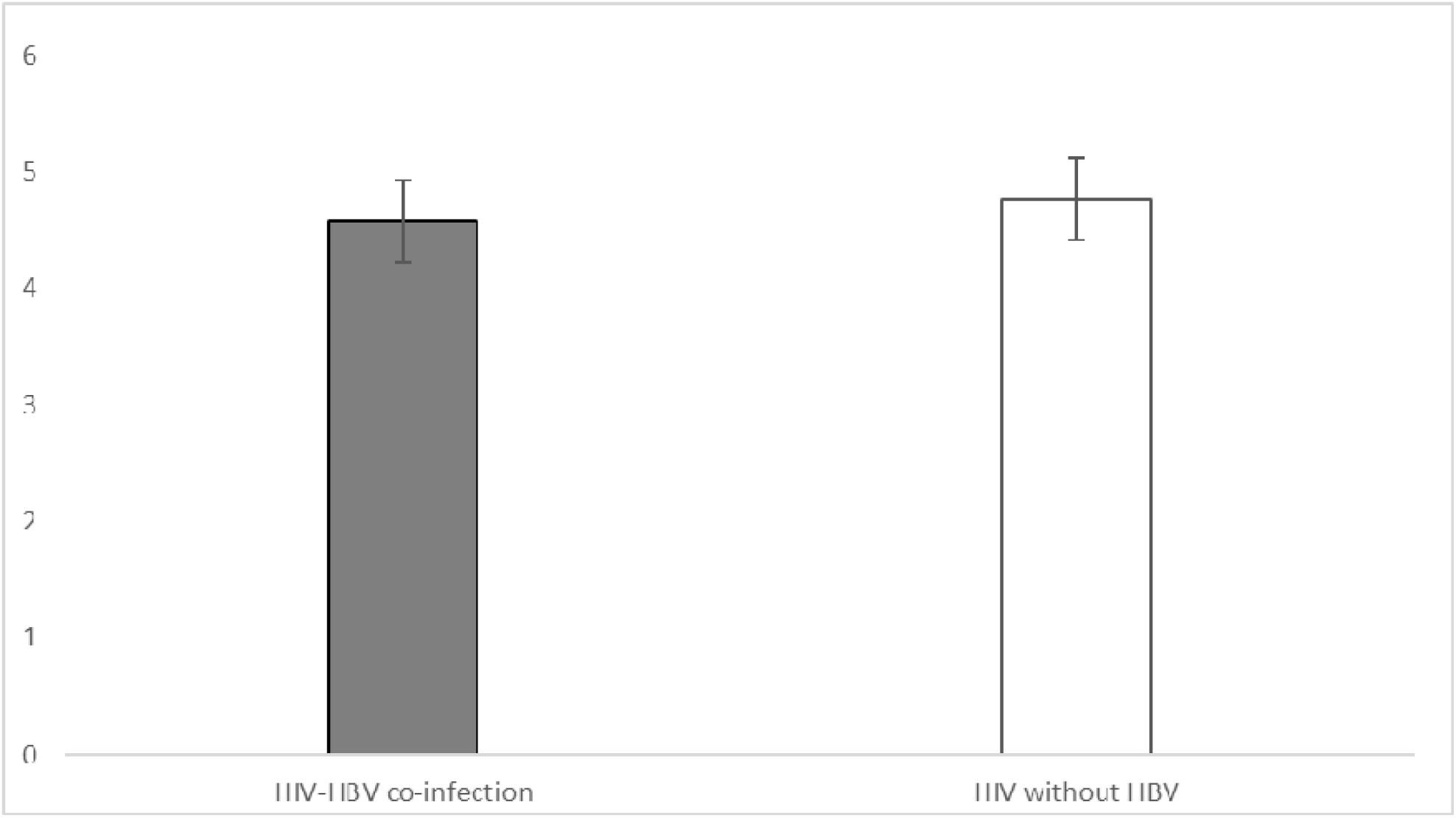
Years of survival/follow up by HIV-HBV co-infection or HIV only status.

Among the HIV-HBV co-infected, 37 (86%) failed to spontaneously clear the HBV infection and the majority of them continued to remain HBV positive over the years till 2020 (figure 5). Only 4 (9.3%), all adults among total of 43 patients were able to spontaneously clear HBV infection during the normal course, anti-HBc antibody was found in all 5/5. Those who could clear HBV were younger and had higher mean CD4 counts as compared to those with HBV persistence as shown in table no. 3. In the initial years, the HAART regimen used was Zidovidine based regimen, and post 2015 according to the new guidelines patients were put on Tenofovir, Efavirenz and Lamivudine based regimen. Prior to 2015 concomitant anti-HBV treatment was administered to only those patients who showed liver derangements.

**Figure 5:**
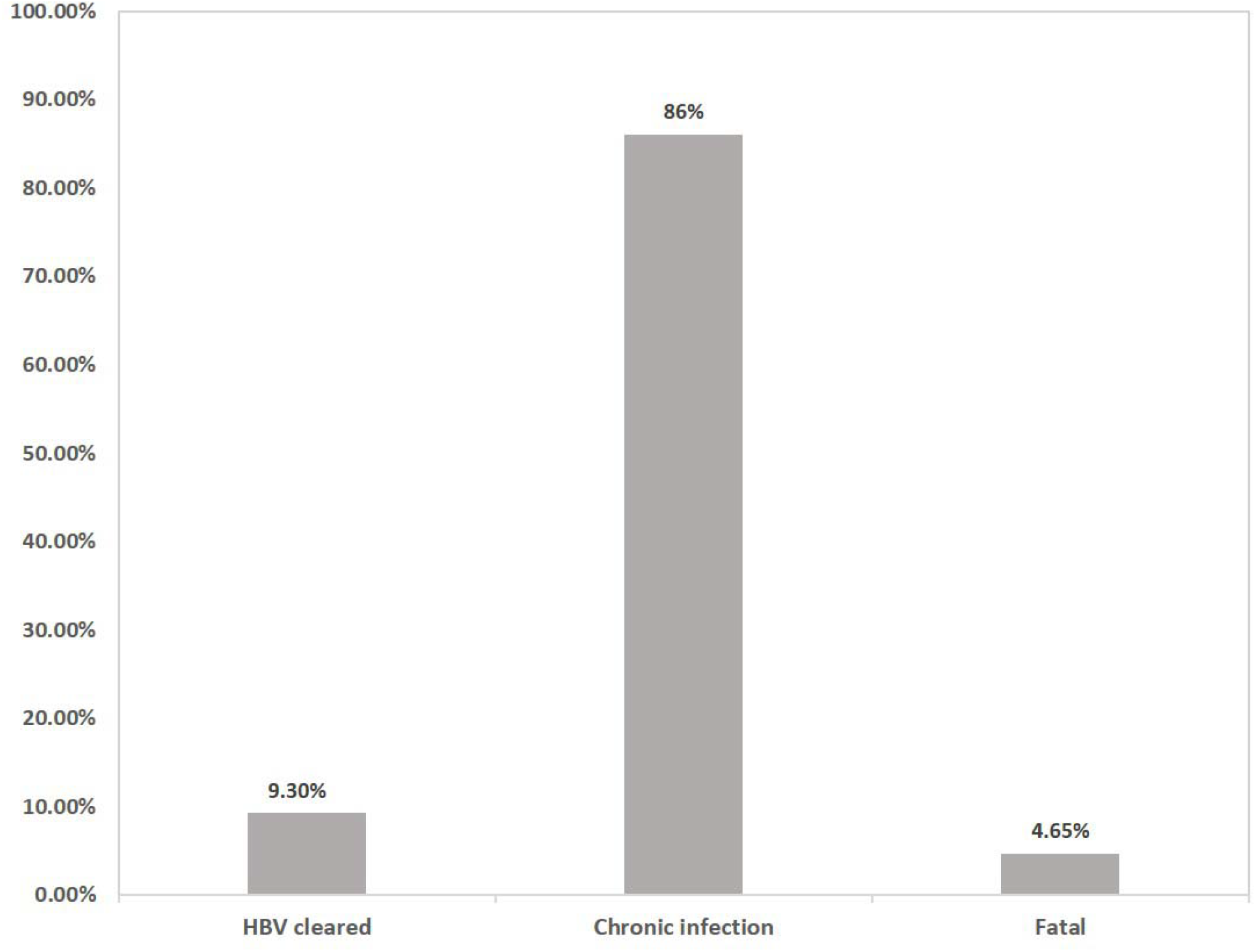
Outcomes and clearance status among the HIV-HBV co-infected.

On statistical analysis after the application of the Chi-square test to the prevalence of co-infection among males versus females, the p-value was found to be 0.92, which is statistically non-significant. Statistical significance by paired T-test for various parameters showed that CD4 values in the co-infected group were higher than HIV only group; this association was found to be significant with a p-value of 0.02, and other baseline parameters did not show any significant association. Comparing the significance of mean age and mean values of CD4 counts amongst those who cleared HBV versus those with HBV persistence, there was no significant difference among them.

## Discussion

Our research has unveiled significant insights, firstly the overall prevalence of Hepatitis B Virus (HBV) among individuals living with HIV was determined to be 2.4%. This rate is similar to studies conducted in North India, sharing a similar transmission route pattern. Somehow, comparatively our findings indicate a slightly lower prevalence (12,13). The seroprevalence findings of our study are also similar to that observed among high-risk populations in other Asian countries like Iran (19). The prevalence of co-infection observed was not only far lower than that reported from studies in western countries having an average prevalence of around 5-6%, but it was also lower than the national average prevalence among healthy populations of 4%. On the other hand, the HBV seroprevalence among the co-infected was similar to that reported among the general population in Central India, thus indicating that the HIV population in our area may not be exposed to any additional risk factors as compared to the general population (10). As for Hepatitis C Virus (HCV), our investigation uncovered a very low seroprevalence. Thus, at present HCV does not appear to pose a significant challenge within the scope of our study.

Various factors and their interplay might be responsible for the observed low seroprevalence. The risk of HIV and HBV co-infection is not only affected by the individual’s age at the time of exposure to both viruses but also by the route of transmission and the vaccination status as well (20). In western countries, HBV is typically acquired during adolescence or early adulthood either through IV drug injections or through sexual activity (12). There has been a declining trend in its prevalence due to increased immunization coverage both in the East and the West, most of our population was unimmunized for HBV. However, since ours is a retrospective study we may be lacking accurate data regarding the immunization status of our patients who registered for HAART in earlier years. However, the effect of immunization is visible in the later part of our study where a decline is seen in the incidence.

Over the years with the advent of immunization policy including HBV vaccination for children, safer and more effective ART, HIV-HBV co-infections showed a downward trajectory with a nadir observed in 2015, with almost a plateau thereafter. We observed a drop in co-infection rates in recent years, it seems to change from 2015 onwards following the effects HBV vaccine inclusion in the universal immunization schedule (21). Increased popularity of single-use syringes and disposable surgical ware over the years could also be a factor. Additionally, it could be due to the change in regimen when Tenofovir, Efavarence, and Lamivudine based regimens were introduced replacing Zidovudine based regimen. However, in such case this observed decline may soon fail to sustain given the possibility of the gradual emergence of drug resistance which is known to arise after a few years of introducing newer therapy in such situations (22).

Age-specific data in our study highlights an important finding that co-infected people are likely to acquire the infection at a younger age in comparison to persons with HIV alone; This discovery aligns with findings from a subset of prior studies (16,23), but is not universally corroborated by all existing research. This underscores the importance of directing attention towards these vulnerable groups at an earlier stage and calls for early targeting and bolstering vaccination coverage among them.

Our study showed that co-infected individuals were more likely to be male, although in our case it was not found to be statistically significant as reported from studies conducted in the West (23). This difference could be due to the difference in transmission routes between western countries, eastern Indian states, and central India. Earlier studies from India mostly reported the seroprevalence of co-infection to be more when transmitted among men who have sex with men or among people injecting IV drugs (13,15). Since the majority of our population had acquired the co-infection through the heterosexual route, this difference in route of transmission appears to be the reason behind the comparatively low prevalence of co-infection observed in our study population.

Earlier studies have reported that the CD4 values of the HIV-HBV co-infected group is somewhat lower than HIV only group; however this was not the case with our population, on the contrary, we observed that the co-infected group had a higher mean CD4 count (8,24). The association was found to be significant with a p-value of 0.02, this might an area of further research to uncover any new associated variables which we might be missing. In our limited cases with HIV-HCV co-infection, they had a chronic infection and a lower CD4 count as compared to HIV only, this is in agreement with earlier studies (25). Other baseline parameters did not show any significant association.

A substantial proportion of our subjects remained HBV positive for more than 2 years, demonstrating the inability of the clearance mechanisms in timely getting rid of the infection, specifically in the subset of cases involving children. Among these cases, none were able to achieve HBV clearance, further accentuating the challenge of HIV-HBV co-infection. In contrast, a minority of the co-infected adults in our study exhibited spontaneous HBV clearance during follow-up testing. These findings emphasize that HIV does interfere with the natural course of HBV infection as found in earlier studies (4,5). Although there is a high rate of spontaneous clearance of HBV (>90%) in immunocompetent adults, the chronic infection has been reported to develop in around 20 percent of adults with HIV infection after exposure to HBV, this percentage is far greater in our cohorts which needs to be deeply understood in our context.

Moreover, this also highlights there is an urgent need to make a policy decision in our country and countries who haven’t started a compulsory HBV vaccination to start this practice in all persons detected with HIV infection without fail, as practiced in the west. The benefits are indisputable since the initial financial occurring therein holds the potential to avert larger costs associated with managing chronic diseases and their ensuing complications.

On carefully examining the disease severity, it was noted that despite not having alarmingly low CD4 counts, the co-infected persons exhibited greater morbidity as revealed by ultrasound performed on co-infected patients. Upon comparing the proportion of unfavorable outcomes within our study population to that of other research endeavors, it was surprising that the mortality rate among our subjects was unexpectedly high. This might be due to the fact that our patients mostly belong to poor socio-economic strata which is the case in Bundelkhand region leading to poor nutritional status, as also evidenced by their low mean body weight (Table 2).

When we compare the mortality among the co-infected versus HBsAg negative group, co-infection resulted in a higher percentage of death, however, it was not found to be of statistical significance. As evident from the follow-up data, there is a gap in longevity between the HBV co-infected and HIV only. The co-infected are likely to die early and this gap in longevity is bound to increase as the follow-up period approaches the 15 to 20-year period when chronic HBV infection is known to cause death due to complications such as cirrhosis and hepatocellular carcinoma (26).

## Conclusion

Despite the low seroprevalence seen in the specific population in our study, the majority of co-infected failed to clear HBV, with a notable percentage showing an adverse outcome. To catalyze a transformative shift in their prognosis, the current situation unequivocally necessitates the prompt implementation of universal HBV vaccination for those living with HIV. Such a strategy holds the potential to grant them an extended life span relatively unburdened by disease. Overall, by focusing on these preventive measures and interventions, we can potentially mitigate the impact of such co-infections, improve public health outcomes, and decrease the long-term burden on healthcare systems.

## Limitations

We lacked vaccination history, natural history before presenting to the ART center for all subjects, these things are often complicated by the fact that HIV has a significant social taboo associated with it, hence people prefer not to disclose past history. Our study was also limited by time constraints, with a maximum follow up of around 10 years. A further follow-up of these cases is required to determine the long-term outcome when most of complications such as cirrhosis and hepatocellular carcinoma are known to appear. Due to the retrospective nature of the study, we could not perform MHC expression studies, although we wanted to find MHC-1 expression rates in our study population which is associated with clearance of infection and compare it to that among the general population (27). Nevertheless, we believe this study is the first of its kind in the Central India and helps to provide important novel insights.

## Conflicts of interest

We declare that there are no conflicts of interests involved. All authors have contributed sufficiently to the research and in writing the manuscript.

## Funding

No external funding was provided for the study, no specific grant from any funding agency in the public, commercial or not-for-profit sectors has been used. The diagnosis and clinical investigations were carried out as part of the routine patient care at the hospital which is partly funded by NACO.

## Data Availability

All data produced in the present study are available upon reasonable request to the authors.

https://naco.gov.in/

## List of abbreviations

ALT: Alanine aminotransferase
ART: Anti-retroviral therapy
AST: Aspartate aminotransferase
ALK: Alkaline phosphatase
CD4: Cluster of differenciation 4
HAART: Highly active anti-retroviral therapy
HbeAg: Hepatitis B envelope antigen
HBcAg: Hepatitis B core antigen
HBV: Hepatis B virus
HCV: Hepatitis C virus
HIV: Human immunodeficiency virus
MHC: Major histocompatibility complex
MSM: Male having sex with male
NACO: National AIDS control organization
PCR: Polymerase chain reaction
S bil: Serum bilirubin

## Notes

### Competing Interest Statement

The authors have declared no competing interest.

### Funding Statement

This study did not receive any funding

### Author Declarations

The review and analysis work was performed after obtaining due approval from the institutional human ethics committee at Bundelkhand Govt. Medical College and Hospital, approval number IECBMC/2020/10.

